# The impact of financial support and budgeting on medication availability and purchasing behavior in the Nigerian primary healthcare system

**DOI:** 10.1101/2024.08.23.24312494

**Authors:** Brittany Hagedorn, Rui Han

## Abstract

Previous work has shown that primary healthcare facilities can benefit from both in-kind support (e.g., medication shipments) as well as increased cash-on-hand to spend to address service readiness gaps. However, there is limited evidence on how facility managers choose to spend available cash or how their decisions to manage their facility budgets are affected by in-kind support.

Economic theory suggests that the optimal allocation of cash resources would depend on the context and constraints to how it can be spent, and expenditures would in turn affect the availability of supplies and medications. We test this theory using regression analysis on data from the Nigeria Service Delivery Indicators for Health (SDI), a health facility survey from twelve states in 2013 that included both hospitals and primary healthcare centers (PHCs).

We find that facilities with financial resources available to them have higher availability of essential medicines, especially if the facility had earmarked some cash for medication expenditures. However, earmarking for other expenditure categories did not have the same effect on medication availability, which indicates that budgeting processes are an important factor in ensuring medication availability. We find that cash support had large effect (p < 0.001) on availability and that in-kind donations had a negative effect on the probability of expenditure of medications. Additionally, we find the difference between hospitals and PHCs is due to their financial situation (variables become insignificant once support variables were in regressions).

Regression analyses also showed that facilities that received in-kind medications had higher availability, but this only had a significant effect in facilities that did not have cash available to spend on medications, implying that facilities are able to address their own supply needs when they have resources available to them. Thus, in-kind supplies should be targeted to facilities that cannot otherwise procure them. Overall, facilities appear to be making effective trade-offs in the context of limited resources and they should receive both cash and support for appropriate budgeting and procurement practices.

## Introduction

Previous studies have shown that incremental financing for primary healthcare facilities can improve structural readiness and service availability in the Nigerian context. This was studied specifically in three Nigerian states through the Nigeria States Health Investment Project (NSHIP), which provided an integrated intervention package that included additional financing direct to the facility, management and budgeting skills training to the officers in charge of facilities, and additional autonomy over facility management and budget allocation. (1)

An important finding from secondary analysis of the NSHIP facility surveys was that both study participation (study arm) and the level of revenues available at the facility level were independently predictive of performance improvements (2). Additional work has shown that facility-level autonomy, management practices, and budget control were also positively predictive of performance. (3)

With this in mind, we hypothesize that it is not just control of a budget that drives performance, but rather how that money is allocated and spent. To approach this question, we utilize an alternative dataset from the Nigerian context that was collected over a similar time horizon as the NSHIP facility surveys. This allows us to investigate whether the positive impact of financing on structural readiness of facilities can be broadly observed outside of high-intensity intervention programs like NSHIP, and the specific effect that budgetary practices have on outcomes.

## Methods

In this analysis, we aim to address two closely linked research questions. **First:** Does an increase in facility-level funding improve structural quality? Second: If so, how does this depend on what the facility’s needs are (e.g., if they already have in-kind donations) and how do limitations on how the money can be spent affect a facility manager’s spending behaviors?

### Conceptual Model

Our conceptual framework follows from the Donabedian model, with expectation that funding levels provide the resources to acquire the ‘inputs’ such as equipment and supplies that are required in order to generate ‘outputs’ as measured by service volumes.

We take an economic viewpoint and assume a rational, empowered facility manager who takes an active role in optimizing their budget allocations to acquire inputs and meet the needs of their facility. In doing so, they presumably spend their ‘first dollar’ on the most essential items for providing health services, such as availability of essential medicines and supplies at the facilities in low-resource settings, thus creating a sub-linear relationship between funding and performance.

We seek to uncover whether there is evidence of this relationship in a context like Nigeria, where autonomy is relatively limited compared to other low-income countries. To do so, we examine if and how cash and in-kind support affect readiness differently, and whether there is evidence that it improves readiness by changing operator behavior (such that they spend on medicines only when they need to).

### Data

For this study, we conduct secondary data analysis on the Nigeria Service Delivery Indicators for Health (SDI), a health facility survey conducted in twelve Nigerian states from July 2013 to January 2014. This is the only Nigeria SDI for health survey publicly released as of June 2024.

We utilize two survey modules. Module 1 includes questions on facility readiness (infrastructure, equipment, medicine and supply availability) and covers 2,385 health facilities across twelve states. Module 4 includes questions on financials (cash support, in-kind non-cash support, and facility expenditures) and covers 1,192 facilities across six states. We use only the facilities that have complete data in both modules, resulting in a survey sample from Bayelsa (175 facilities), Imo (218), Kaduna (205), Kogi (203), Osun (203) and Taraba (188). These are split across 145 health posts, 788 health centers and clinics, and 259 hospitals.

### Variables

For our analysis, we construct an indicator from the facility survey to summarize supply availability into a dependent variable. For each of the thirty essential medicines asked about in the survey, we assign it a score between zero and one, depending on its level of availability:

- 1 = available and non-expired
- ½ = available but expired
- ¼ = sometimes available but not currently
- 0 = never available

These are then averaged, such that the supply availability index is the average score across all thirty essential medicines.

Independent variables include controls (state, facility level, and geography) as well as three constructed variables for the type of budgetary support and practices reported by the facility. The first binary variable indicates whether or not the facility received any kind of in-kind donation of medications. The second categorical variable indicates whether or not the facility received any kind of cash support and if so, whether it was earmarked at all or for medicines specifically. The third binary variable indicates whether a facility reported cash support from more than one source.

### Relationship between support and supply availability

Our first analysis examines the relationship between cash and non-cash support reported by the facilities and their performance on the structural quality indices described above. We use linear regression to test this hypothesis and report coefficient estimates and p-value significance. Further, we compare a null model (controls only) to two alternative formulations of the regression including the support variables, using ANOVA to test for statistical significance.

### Relationship between earmarking and expenditure behavior

Our second analysis examines whether the facility manager behaves as a rational actor, more likely to spend money on medications when there is budget earmarked for this, and when they don’t have an alternative source of products (i.e., they would spend on medications when they especially need to). We use logistic regression to test this hypothesis and again compare a null model (controls only) to regressions including the relevant variables, using ANOVA to test for significance.

## Results

### Survey data descriptive statistics

Facilities were distributed across six states, with most falling into the health center category. Taraba is the exception, with the majority of facilities being health posts, the lowest level of facility. Of all facilities, 18% had no cash support, 20% had cash support that was not earmarked, 14% had cash earmarked for budget categories other than medications, and 48% had cash support earmarked for medication. (Table 1)

**Table 1.** Number of facilities in the available survey sample, by type. Cash, no earmark = the facility received cash support, but it was not designated for any specific use. Cash, earmark other = the facility received cash support, and it was earmarked for an expenditure other than medications. Cash, earmark meds = the facility received cash support, and it was earmarked to be used to purchase medications.

| State | Geography |  |  | Facility type |  |  | Budget/Earmarking |  |  |  |
| --- | --- | --- | --- | --- | --- | --- | --- | --- | --- | --- |
|  | Rural | Urban | Semi-urban | Health post | Health center | Hospital | No cash support | Cash, no earmark | Cash, earmark other | Cash, earmark meds |
| Bayelsa | 101 | 21 | 53 | 3 | 136 | 36 | 42 | 14 | 11 | 108 |
| Imo | 190 | 25 | 2 | 23 | 147 | 47 | 21 | 90 | 20 | 86 |
| Kaduna | 131 | 30 | 43 | 8 | 149 | 48 | 28 | 9 | 15 | 153 |
| Kogi | 113 | 24 | 65 | 6 | 141 | 55 | 24 | 90 | 22 | 66 |
| Osun | 44 | 85 | 74 | 17 | 150 | 36 | 86 | 5 | 84 | 28 |
| Taraba | 161 | 18 | 9 | 88 | 65 | 35 | 17 | 28 | 12 | 131 |

Across all facilities, medication availability varied widely for the various products, from 13% (azithromycin) to 83% (Folic acid) fully in-stock. Partial availability results of in stock but expired (score of ½) and sometimes available but not today (score of ¼) were present in a meaningful but smaller portion of facilities, indicating that it is less common to have occasional availability of a product. These patterns of availability were generally consistent across the six states included in this analysis, with facilities from Taraba reporting partial availability somewhat more often than in other states. (Figure 1)

**Figure 1.**
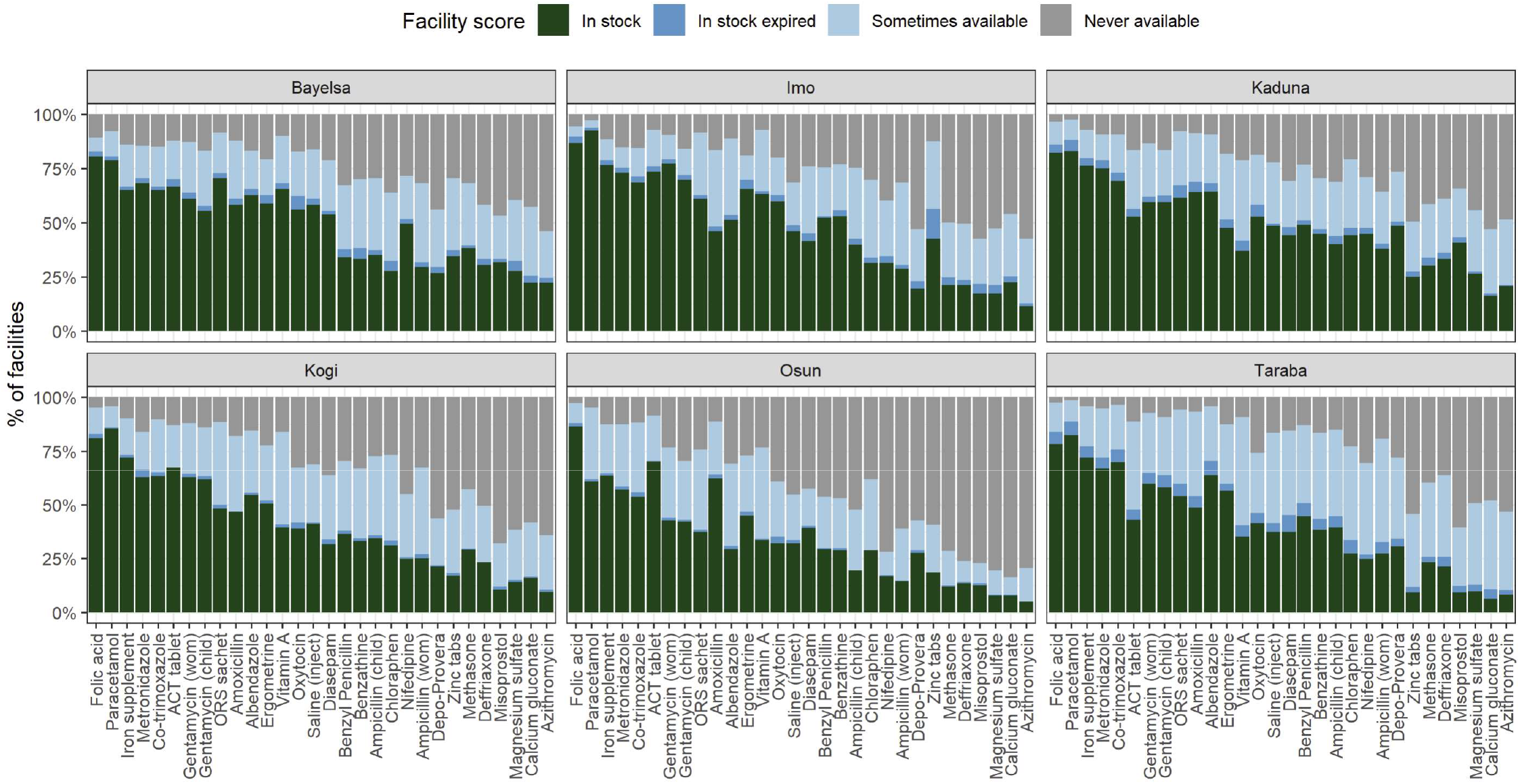
Product availability across all facilities, by state. Bar height represents the proportion of facilities with a particular survey result for that product. The distinction between sometimes and never available responses was reported by the interviewee and not verified, assumed to be truthfully and accurately reported.

There was also variability in performance between facilities of different levels, with hospitals generally having the highest scores, followed by health centers, and health posts the lowest scores. This was true across states, with the smallest differences observed in Taraba (average scores of 0.63, 0.58 and 0.41 respectively) and largest in Bayelsa (average scores of 0.74, 0.52 and 0.06 respectively). (Figure 2)

**Figure 2.**
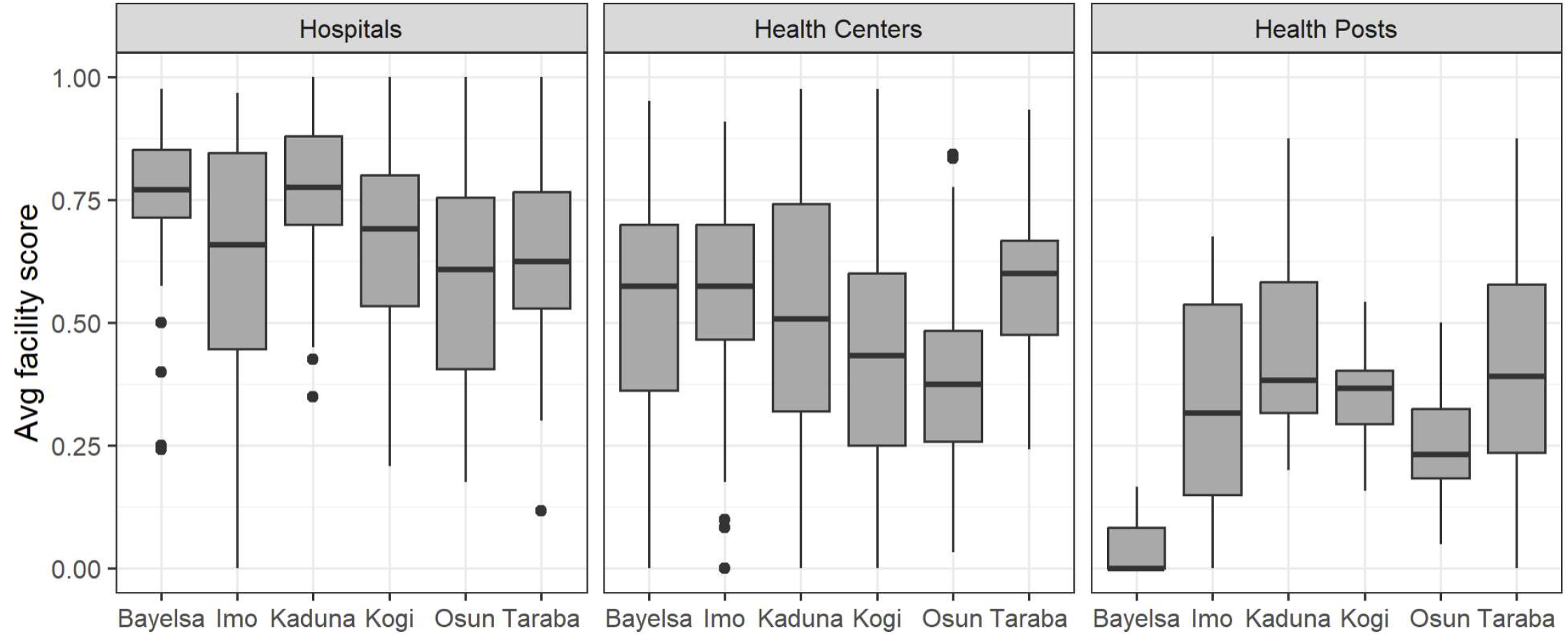
Average scores per facility, by state and facility level. Values represent average scores across all thirty essential medicines. Sample excludes facilities with unknown level (NA), which was 2.8% of the facilities in the six states. Boxplot values: center line = median, box height = 25^th^/75^th^ percentiles, whiskers = 5^th^/95^th^ percentiles, dots = outliers.

There were a variety of cash and in-kind (non-cash) support arrangements that affected facilities’ resources available to invest in medication availability. Non-cash support for any type of items was received by 84.2% of facilities with medicines received by 61.3% of facilities, and cash support from any source was received by 81.7% of facilities. Among those that received cash support, they received this from an average of 1.3 sources (75^th^: 2, max: 6). Among those that received non-cash support, they received this for an average of 3.4 types of items (75^th^: 5, max: 18). These groups are further broken down in Table 2.

**Table 2.**
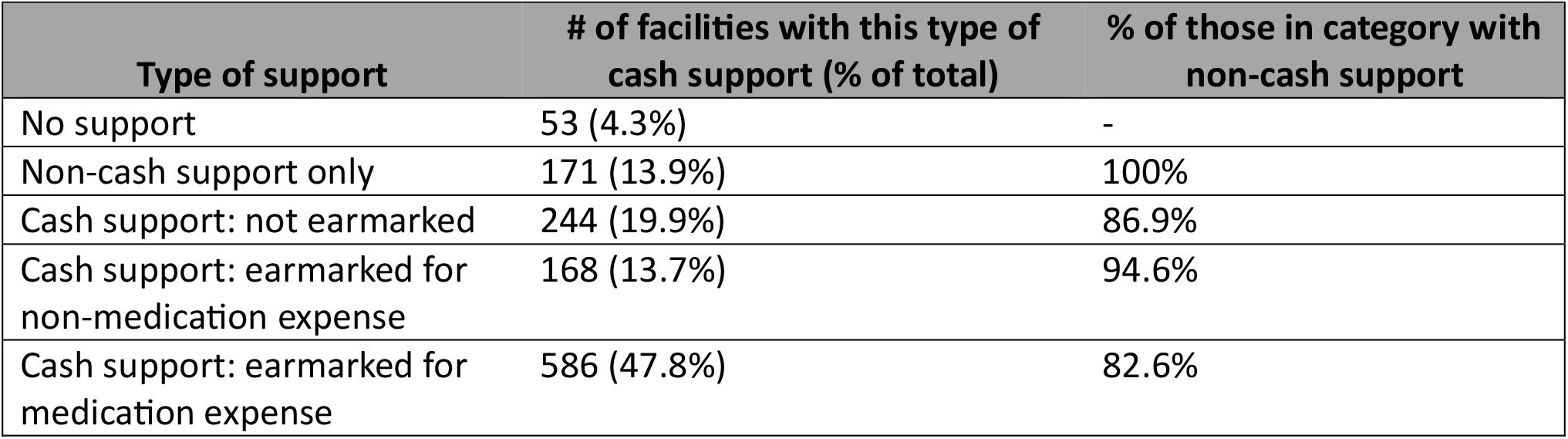
Breakdown of facilities by level of support, cash and non-cash. Calculations exclude NA values, which were less than 1% of facilities in the survey.

| Type of support | # of facilities with this type of cash support (% of total) | % of those in category with non-cash support |
| --- | --- | --- |
| No support | 53 (4.3%) | - |
| Non-cash support only | 171 (13.9%) | 100% |
| Cash support: not earmarked | 244 (19.9%) | 86.9% |
| Cash support: earmarked for non-medication expense | 168 (13.7%) | 94.6% |
| Cash support: earmarked for medication expense | 586 (47.8%) | 82.6% |

To validate that the data is internally consistent and of reasonable quality for the analysis, we compared facility reporting of a) whether a facility received cash support and b) whether they listed cash expenditures. In doing so, we did not find evidence of misreporting, as facilities that received no cash support had higher rates (94% compared to 49% for facilities receiving cash support with no earmark, 78% for facilities receiving cash support earmarked for non-medication expenditures, and 6.5% for facilities receiving cash support earmarked for medication purchases) of claiming that they had no medication-expenditures across any categories, suggesting the presence of budget constraints when there was not an external provision of cash.

### Regression Results

We examined the impact that facility support has on supply availability by running three regressions and used ANOVA to compare whether the models performed better with the addition of the support category variables. The baseline model included only three categorical variables (geography, facility type, and state) and all three were statistically significant, resulting in an adjusted R2 value of 0.21. Hospitals are associated with better medicines readiness compared to health centers, while health posts are associated with worse medicines readiness relative to health centers. A facility in urban areas is associated with better medicines readiness relative to a facility in rural areas. The second model included cash support type and whether there were multiple sources, which were both significant at p < 0.001. This increased the adjusted R2 to 0.277 and was a significant improvement. Comparing the coefficient estimates of the baseline model and the model with financing variables, the coefficient estimates on hospitals shrank by 19% and the coefficient estimate on health posts shrank by 15%. Finally, the third model also included a binary variable for in-kind support in the form of medication donations and the interaction term with cash support. In-kind support was significant at p <0.05 but the interaction term was not; though the adjusted R2 increased slightly, it was not a significant improvement to the model. (Table 3)

**Table 3.** Regression results for analysis on the effect of cash and in-kind support on medication availability. Outcome metric was facility-level average medication availability score for the thirty essential medicines reported in the survey. Cash support = categorical variable with four levels, indicating whether the facility reported receiving cash support from an external source and if so, whether and how it was earmarked for specific expenditures categories. Earmark for other = cash support is earmarked only for non-medication expenditures. Earmark for meds = cash support is at least in part earmarked for medication purchases. Multiple sources = binary variable indicating whether the facility reported receiving cash support from more than one external source. In-kind support = binary variable indicating whether the facility reported receiving any direct donations of medications.

| Variable (default) | Baseline model<br>Coefficient / p-value |  | Consider cash support<br>Coefficient / p-value |  | Consider all support<br>Coefficient / p-value |  |
| --- | --- | --- | --- | --- | --- | --- |
| Geography (rural) |  |  |  |  |  |  |
| Semi-urban | 0.03 | 0.10 | 0.03 | 0.12 | 0.028 | 0.11 |
| Urban | 0.05 | <0.01 | 0.06 | <0.001 | 0.057 | <0.001 |
| Type (health center) |  |  |  |  |  |  |
| Hospital | 0.17 | <0.001 | 0.14 | <0.001 | 0.14 | <0.001 |
| Health post | -0.15 | <0.001 | -0.12 | <0.001 | -0.12 | <0.001 |
| State (Bayelsa) |  |  |  |  |  |  |
| Imo | 0.03 | 0.14 | 0.04 | 0.10 | 0.03 | 0.22 |
| Kaduna | 0.02 | 0.43 | -0.004 | 0.84 | -0.009 | 0.66 |
| Kogi | -0.08 | <0.001 | -0.078 | <0.001 | -0.089 | <0.001 |
| Osun | -0.15 | <0.001 | -0.093 | <0.001 | -0.134 | <0.001 |
| Taraba | 0.04 | 0.09 | 0.01 | 0.68 | -0.005 | 0.82 |
| Cash support (no cash) |  |  |  |  |  |  |
| no earmark | - | - | 0.10 | <0.001 | 0.13 | <0.001 |
| earmark for other | - | - | 0.09 | <0.001 | 0.02 | 0.71 |
| earmark for meds | - | - | 0.16 | <0.001 | 0.18 | <0.001 |
| Multiple sources (none) | - | - | 0.05 | <0.001 | 0.05 | <0.01 |
| In-kind support (none) | - | - | - | - | 0.07 | 0.02 |
| Interaction term |  |  |  |  |  |  |
| In-kind * no earmark | - | - | - | - | -0.06 | 0.14 |
| In-kind * earmark other | - | - | - | - | 0.07 | 0.15 |
| In-kind * earmark meds | - | - | - | - | -0.05 | 0.14 |
| Adjusted R2 | 0.21 |  | 0.277 |  | 0.284 |  |
| ANOVA | - |  | p-value < 0.001 (vs. Baseline) |  | p-value <0.01 (vs. cash support) |  |

We then examined whether there was evidence of change in expenditure behavior in response to the types of cash and in-kind support that they received, using a logistic regression to predict the likelihood that a facility would spend on purchasing essential medicines. Comparing the regression models showed that including the variables for types of cash and in-kind support increased the pseudo R2 value from 0.30 to 0.68, a significant improvement. All included support variables were significant, with the cash support categorical variable at p <0.001 and in-kind support at p = 0.01. The odds ratios were substantial positive, with facilities with cash available to spend on medications seeing a 10x or more increase in the probability of spending on medications. Simultaneously, the odds ratio for in-kind support is below one, indicating a nearly 40% reduction in the probability of spending on medications. Further, the facility type was no longer significant when considering the type of support variables. (Table 4)

**Table 4.**
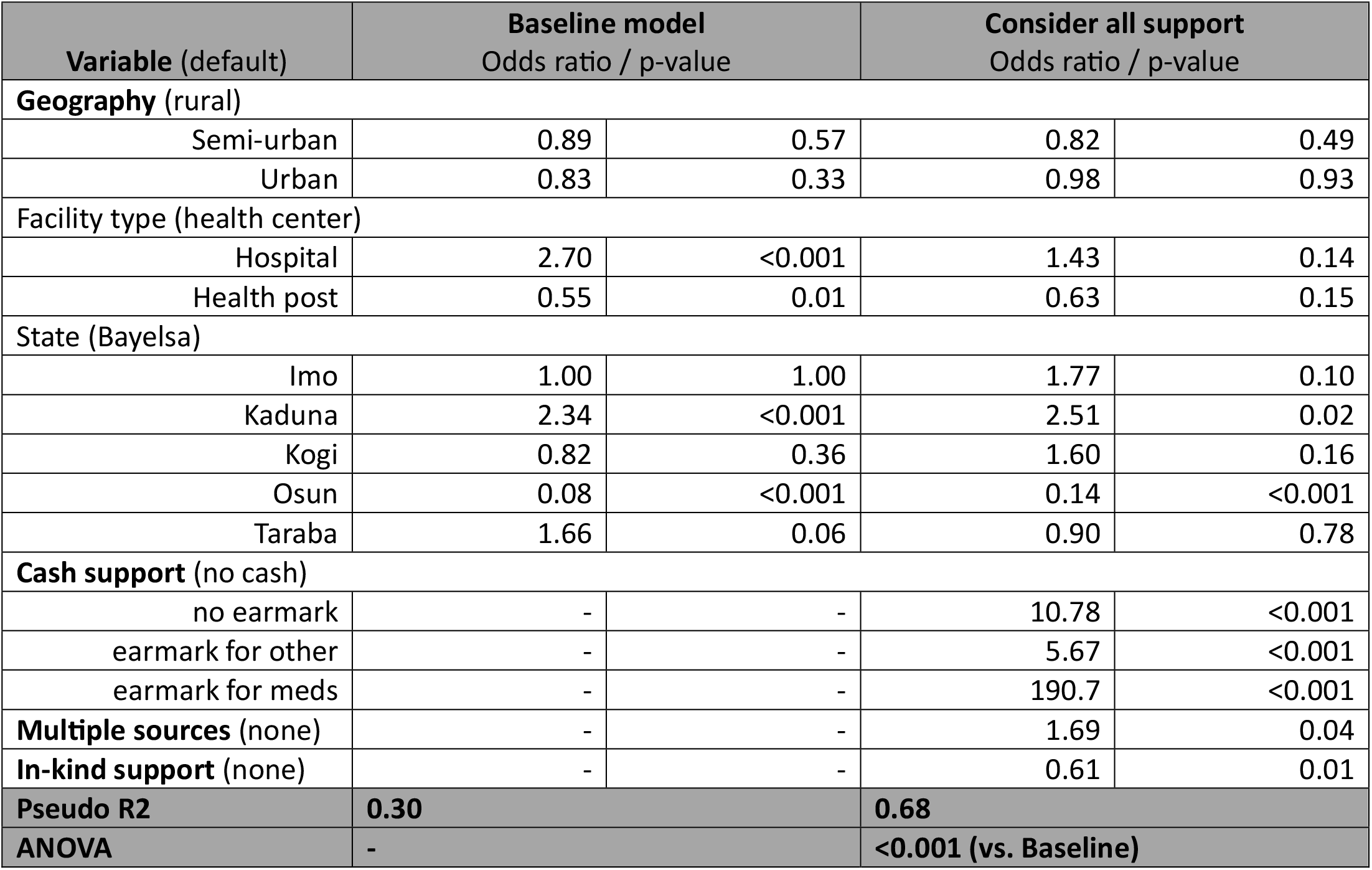
Regression results for analysis on the effect of cash and in-kind support on expenditure behaviors. Outcome metric was log odds of the facility-reported expenditure on medications. Calc. coefficient = calculated coefficient value for the average facility. Cash support = categorical variable with four levels, indicating whether the facility reported receiving cash support from an external source and if so, whether and how it was earmarked for specific expenditures categories. Earmark for other = cash support is earmarked only for non-medication expenditures. Earmark for meds = cash support is at least in part earmarked for medication purchases. Multiple sources = binary variable indicating whether the facility reported receiving cash support from more than one external source. In-kind support = binary variable indicating whether the facility reported receiving any direct donations of medications.

For comparison, the average facility that received no cash support had an estimated 6% probability of purchasing medications, whereas a facility with cash that was unearmarked had a probability of 51% and a facility with cash earmarked specifically for medication purchases had a probability of 93.5%.

We also performed these sets of regression analyses looking only at the subset of facilities that are health centers, which was 66% of the facility sample. Further, we conducted a sensitivity analysis on the structure of the performance score, removing the use of partial values to represent occasional availability. The results were qualitatively similar to those described above and are not reported here.

## Discussion

Let’s assume a facility operator manages a PHC facility. The operator has a budget and procures and allocates medicines, supplies, labor, and other inputs for service delivery. The operator has the objective to maximize the readiness of the facility to deliver the service package, and in doing so, we would expect three things to occur. First, with additional financial resources available thanks to cash support, they should spend in part on increasing the number of medications in stock. Second, when the money is earmarked for medications, it should be spent that way and improve medication availability. Third, when the facility has alternative ways to obtain medications, such as in-kind donations, they should reallocate some of their financial resources to other needs, thus reducing the probability of spending directly on medications.

In these analyses, we find evidence supporting all three of these effects. Further, we find that the type of cash and in-kind support that facilities receive affects medication availability and purchasing choices in different ways depending on the context. Both cash support and in-kind medication donations were positively associated with medication availability at the facility level, but cash support had a larger effect.

In-kind donations increased availability by 7% in facilities without any cash support and 14% in facilities with cash support that was earmarked for non-medication purchases, but by only 1-2% in facilities with the resources to purchase their own medications. This suggests that when facilities can purchase their own medications, in-stock donations do not have substantial value.

In comparison, facilities with unrestricted cash had 13% higher availability and those with cash specifically earmarked for medication purchases had 18% higher availability. Considering that these coefficients apply to a scale of up to thirty essential medicines, this equates to 3.9 and 5.4 additional medicines in-stock, respectively, from these types of cash support.

Taken together, this suggests that facility managers were spending available financial resources to increase the availability of essential medicines and provide better healthcare services. However, when products are donated directly to the facilities, or the cash is not restricted, managers are making trade-offs and spending on other priorities.

We observe that in the baseline model for medication availability, the type of facility was highly significant; however, once we control for financial resources, ability to purchase drugs, and in-kind support, the effect of facility type is modestly reduced, which means that hospitals outperform PHCs in part because they have the resources to purchase drugs.

The interaction term between cash support and in-kind medication donations provides some interesting insights. While it is not significant, possibly due to sample sizes as small as 26 in these sub-categories. That said, it is interesting to note that for facilities with both in-kind support and funding earmarked for non-medication, the interaction term coefficient was positive, while the others were negative. This implies that in-kind support has a positive effect for facilities that don’t otherwise have resources to spend on medications (net 14% improvement). In contrast, in facilities that had funds available that could be spent on medications, the net effect of in-kind support is net only 1-2% improvement. From this, we conclude that in-kind donations should be targeted only where they are most useful.

When it comes to predicting expenditure behaviors, facilities made choices that demonstrate that they are effectively making trade-offs in the context of limited resources. We conclude this from the combination of a) cash support had large and statistically significant (p < 0.001) effects and b) in-kind donations had a negative effect of the probability of expenditure of medications. Additionally, we find that the difference between facility types is due to their financial situation and not inherent to their level, based on the fact that the significance of these variables disappears once support variables were included in the regressions.

Take together, the two sets of regressions results show that facility managers can be trusted with modest amounts of direct cash support to spend wisely to maximize healthcare service availability and potential for quality of care. This is seen in the positive and sizeable relationship between cash available for spending on medications, expenditure behavior, and medication availability. Further, the effect of in-kind medication support is most substantial in facilities without funding readily available to spend on medicines, suggesting that these types of donations should be targeted to facilities with the least ability to procure.

There are some limitations to this study, mostly stemming from the format of the data that was available to work with. First, the analysis is limited by the fact that it is cross-sectional, which means that we cannot directly draw causal conclusions. This could be addressed if follow-up SDI surveys become available for Nigeria, to allow for direct comparison over time. However, economic theory suggests that it should represent the reality of how incremental funds are spent and that these behaviors are relatively stable over time, so conclusions drawn from this survey should be robust. Second, the potential to detect an effect was hampered by the fact that the publicly available data had been scrubbed and the numeric values for cash and medication support amounts were removed from the survey data (although they were collected), leaving only binary indicators for whether the support was reported or not. This means that some of the conclusions may be confounded if, for example, facilities with higher revenues were more likely to earmark funds for specific purposes. Last, medication and cash support category and not fully independent variables, although we treat them as such in the analysis. In particular, facilities that had cash support earmarked for other purposes were also most likely to receive medicine support, although this would have reduced the likelihood that we would detect a result and so our results are a conservative estimate of effect.

While we understand the need for anonymization, given that the data is quite old, we would recommend a change in data sharing policies. Specifically, when data is adequately old and thus the risk for retaliation or other consequences is gone, the full datasets, including numeric values for revenues and expenditures that were collected, be made publicly available for use in research.

We also recommend two extensions to this analysis. First, additional research should be conducted replicating this analysis with Nigeria’s follow-up SDI data once it is released, as well as in other geographies to test for robustness. Second, expanded data collection to understand the budgeting process itself, who makes earmarking and expenditure decisions, and the eligibility requirements for cash and in-kind support would have been helpful for this analysis.

In conclusion, this evidence shows that when facilities have financial resources, they can and do spend it to ensure medication availability, even without explicit incentives since none were present in this study. Certainly, facility managers may benefit from training and supportive supervision to ensure accountability and appropriate budgeting and earmarking decisions. Additionally, in-kind donations may not be the optimal way to maximize impact at the facility level, at least for facilities that are reasonably well-managed. Last, policies that encourage good budgeting processes, including earmarking at least some cash support for medications, can be an effective strategy for ensuring availability at the facility level.

## Data Availability

All data utilized in this analysis come from the publicly available Nigeria Service Delivery Indicators for Health (SDI) from 2013/2014.

https://microdata.worldbank.org/index.php/catalog/2559

