## Supplement for "The impact of financial support and budgeting on medication availability and purchasing behavior in the Nigerian primary healthcare system"

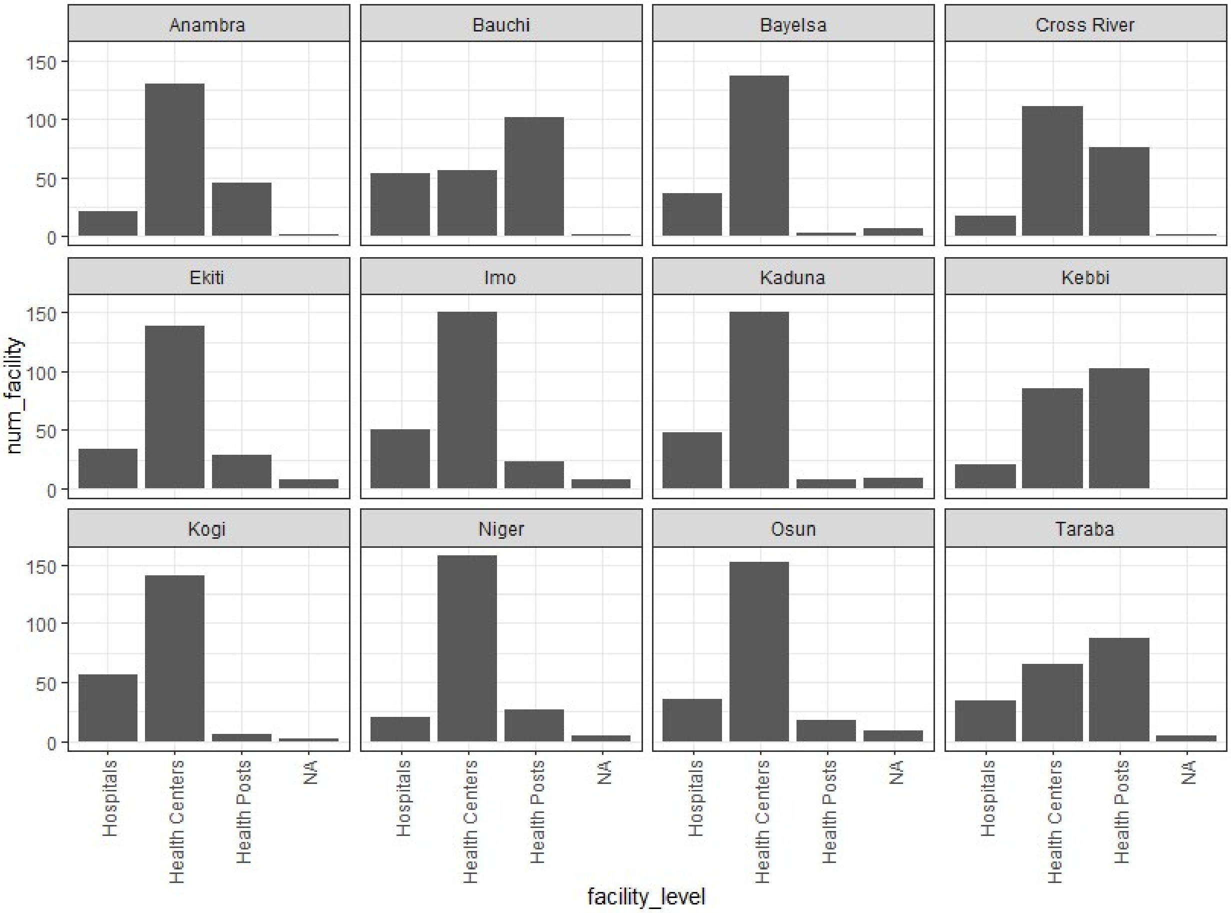


**Figure S1**. Distribution of facilities by type and state. The height of the bar represents the number of facilities falling into a given category, by state. A limited number of facilities did not have a reported facility type in the dataset and are listed under NA.

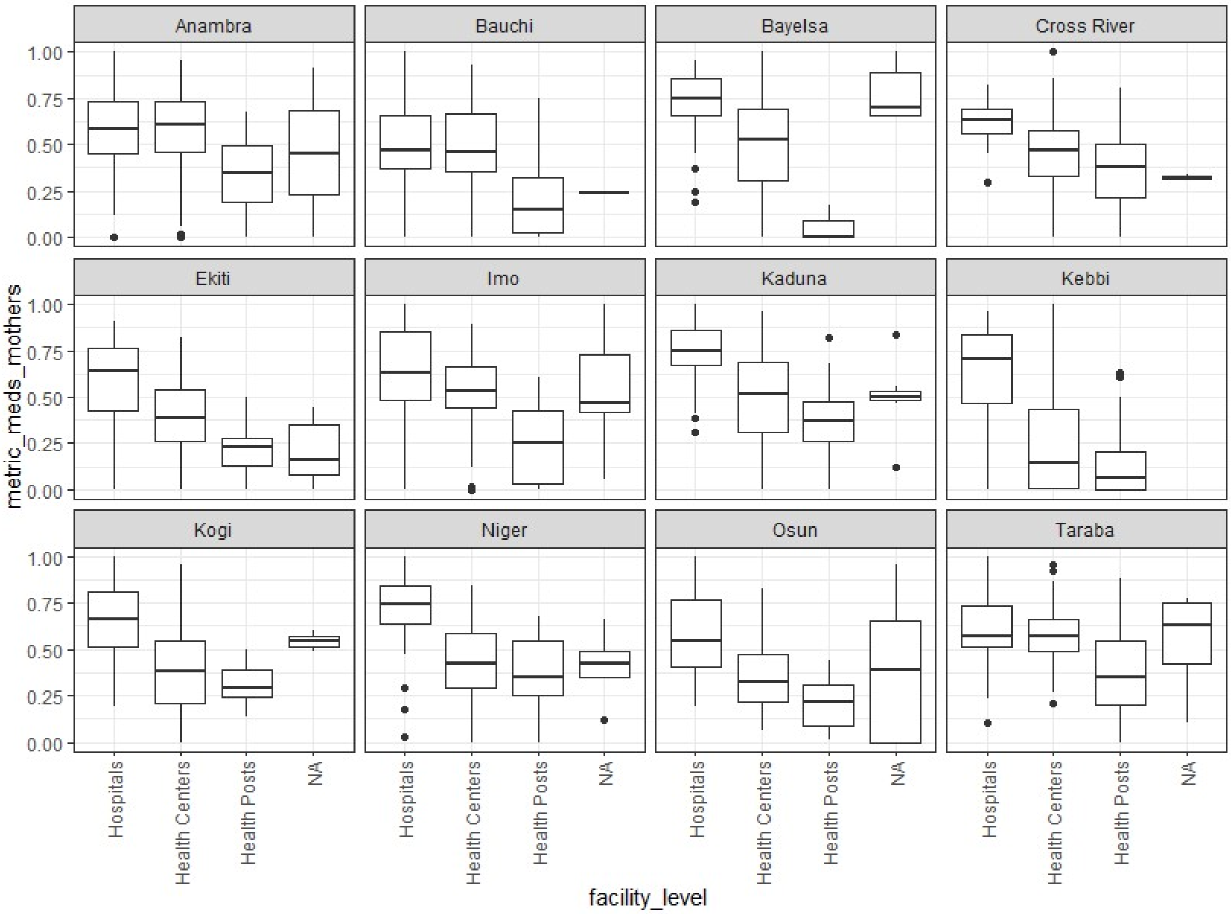


**Figure S2**. Average medication availability scores for essential medicines relevant for maternal care, by facility type and state. The y-axis represents the percentage of the essential medicines that were available in a facility on the day of the survey. Boxplots are read as: horizontal line is the median, rectangle top and bottom are the 25th and 75th percentiles, vertical lines extend to the 95th and 5th percentiles, dots represent outlier values. Facilities that did not have a facility type listed in the survey are summarized in the NA category.
